# Causality or confounding? Applying E values to examine associations between ultra-processed food consumption and weight gain

**DOI:** 10.1101/2024.03.11.24304100

**Authors:** Eric Robinson, Andrew Jones

## Abstract

**Background:** Ultra-processed food (UPF) consumption is associated prospectively with weight gain and obesity in observational studies of adults. Unaccounted for confounding is a risk when attempting to make causal inference from observational studies. Limited research has examined how feasible it is that unmeasured confounding may explain associations between UPF consumption and weight gain in observational research

**Methods:** We introduce the E value to obesity researchers. The E value is defined as the minimum strength of association that one or more unaccounted for confounding variables would need to have with an exposure (UPF consumption) and outcome (e.g., weight gain) to explain the association between the exposure and outcome of interest. We meta-analysed prospective studies on the association between UPF consumption and weight gain in adults. Next, we applied the E value approach and illustrated the potential role that unmeasured or hypothetical residual confounding variables could have in explaining associations.

**Results:** Higher consumption of UPFs was associated with increased weight gain in meta-analysis (RR=1.14). The corresponding E value = 1.55, indicating that unaccounted for confounding variables with small-to-moderate sized associations with UPF consumption and weight gain (e.g., depressive symptoms, trait overeating tendencies, access to healthy and nutritious food) could individually or collectively account for observed associations between UPF consumption and weight gain.

**Conclusions:** Unaccounted for confounding could plausibly explain the prospective association between UPF consumption and weight gain in adults. High quality observational research controlling for potential confounders and evidence from study types devoid of confounding are now needed.

## Introduction

There is significant interest in the potential causal impact that ultra-processed food (UPF) consumption has on health. Observational studies have identified that higher UPF consumption tends to be associated with worse health outcomes, such as weight gain and obesity (1, 2). Prospective studies on this topic are particularly importance as they better allow for inferences on temporal order of associations.

A major challenge in all observational research are unmeasured confounding variables. In line with this, confounding (e.g., residual confounding by social class or lifestyle behaviours) has been discussed as a limitation in numerous studies and reviews on UPFs and health (3–5). To date, there have been limited attempts to quantify how feasible it is that unmeasured confounding could in part attenuate or fully explain prospective relationships between UPF consumption and health outcomes, such as weight gain. A rare exception is a negative control outcome analysis, which found some evidence to suggest that confounding could explain why UPF consumption and cancer were associated in a prospective observational study (4).

In the present article we introduce a recently developed analysis approach to obesity researchers - the E value (6) – to estimate the plausibility that unmeasured confounding could explain observational findings linking UPF consumption with weight gain. The E value is defined as the minimum strength of association that one or more (combined) unaccounted for confounding variables would need to have with an exposure (UPF consumption) and outcome (e.g., weight gain) to explain the association between the exposure and outcome of interest.

When an outcome is predicted using a risk ratio (RR), the E value is calculated as:

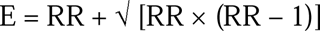

For outcomes predicted using odds ratios and hazard ratios the same equation is used, although some adjustments are made, based on how common the outcome is (e.g. > 15% of individuals have an outcome at the end of follow-up).

Specific E values should not be considered as generically ‘likely’ vs. ‘unlikely’ evidence of confounding potentially explaining exposure-outcome observations, as inference should be based on a case-by-case basis. For example, Gaster et al.(7) conducted a meta-analysis on the association between anti-depressant use and miscarriage risk, concluding that risk of miscarriage was higher among anti-depressant users (RR=1.41). For this RR, the E value = 2.17. Alcohol use was considered as a potential confounder because pregnant women who use anti-depressants are at much higher risk of excessive alcohol consumption than pregnant women who do not. The authors went on to conclude that alcohol use could explain the association between anti-depressant use and miscarriage risk because the relationship (expressed as a risk ratio) between anti-depressant use and alcohol, and alcohol and miscarriage risk are both known to be > 2.17. Yet, it is important to note that the strengths of the association between confounder and exposure and confounder and outcome do not both need to exceed an E-value to provide statistical evidence of potential ‘total’ confounding.

Used alongside the E-value, is the joint bounding factor, B:

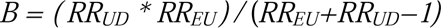

RR_UD_ is the size of association between the potential confounder and outcome and RR_EU_ is the size of association between the potential confounder and exposure. To explore how combinations of confounder exposure and outcome relationships could combine to create statistical conditions for ‘total’ confounding, one sets B (bounding factor) to the E value. In simple language, if the likely size of association between the potential confounder and outcome (or exposure) is larger than E value but the potential confounder and exposure (or out outcome) association is smaller than the E value, the two may still combine to be equal to or exceed the E value and therefore contribute to ‘total’ confounding. For instance, if an E value is 2.00 (RR ∼ 1.33), a stronger association between confounder and outcome (RR = 3.00) and a weaker association between confounder and exposure could exist (RRs > 1.60) to produce ‘total’ confounding and explain away the effect (see Figure 1).

**Figure 1:**
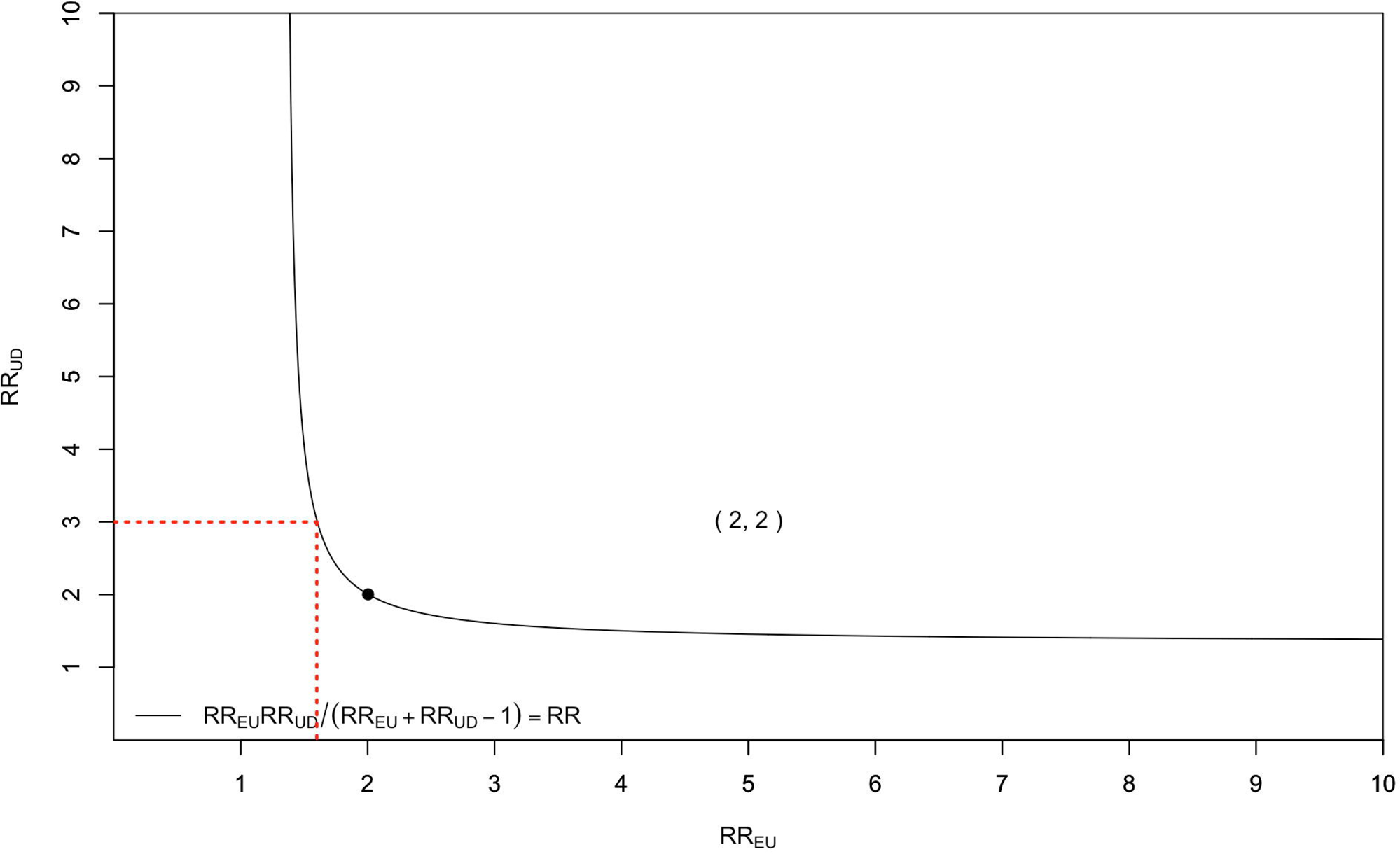
Visualisation of the Bounding Factor. Red dotted lines show example associations between confounder and outcome (RR_UD_) and confounder and exposure (RR_UE_) needed to explain away an effect of RR=1.33. Number in brackets is E Value.

This equation can also be used to estimate by how much (% of effect) combinations of known RR_UD_ and RR_EU_ could account for an observed effect (RR). B = the observed RR, would indicate combinations of RR_UD_ and RR_EU_ could feasibly create ‘total’ confounding and explain away the observed effect. B = 50% of observed RR, would indicate that combinations of RR_UD_ and RR_EU_ could feasibly explain half of the size of the observed effect (‘partial’ confounding).

Here, we introduce the application of the E-value approach in estimating the potential role of confounding in the association between UPF consumption and weight gain, providing an illustrative example of how the E value approach can be used, as well as its limitations, in the context of obesity research.

### UPF consumption and weight gain: identifying potential confounders

Higher UPF consumption has been identified as a potential causal contributor to weight gain and obesity on the basis of observational research. Because both UPF consumption and obesity have sociodemographic and personal characteristic patterning, variables such as age, gender, social class, physical activity and smoking status are typical control variables in study analyses (2, 8) due to concerns over potential confounding. Other potential confounders could be unmeasured. In the present analyses we consider a person’s trait tendency to overeat and experience depression symptoms, as two examples of ‘unmeasured’ confounders.

Socioeconomic status (SES) is a particularly important control variable in diet and health studies. Standard SES measures like education level may not adequately capture the various ways by which social class could indirectly contribute to both UPF consumption and weight gain, resulting in ‘residual’ confounding(9). Residual confounding is typically very difficult to measure, but could be relevant to diet because low SES greatly increases likelihood of decreased access to healthy nutritious food, also known as food insecurity(10). SES measures are somewhat associated with food insecurity(10), but unlikely to capture the negative consequences of food insecurity, resulting in residual risk. Here we therefore treat food insecurity as a quantifiable example of ‘residual’ confounding.

#### Analyses

We identified prospective studies examining UPF consumption and change in body weight among adults from two recent systematic and one recent narrative review on the topic (1, 11, 12). Five prospective studies were identified and meta-analysed. We focused on study effect estimates from analyses relating to weight gain from baseline. See Table 1. We extracted results from models that allowed for prospective weight gain to be accurately quantified and adjusted for the most comprehensive collection of potential confounders, which included demographics (social class, age, sex) and personal characteristics (physical activity, sleep, smoking status) across studies.

**Table 1.** Prospective studies included in meta-analysis.

| *Study* | *Sample* | *UPF characterisation* | *Outcome* |
| --- | --- | --- | --- |
| Mendonca(14) | Normal weight Spanish adults | Quartiles of UPF servings per day<br>FFQ measurement | Risk of developing overweight or obesity<br>8.9 year median follow-up |
| Canhada(2) | Brazilian adults | Quartiles of % total daily energy intake from UPF<br>FFQ measurement | Risk of large weight gain (>1/69kg per year)<br>3.8 year median follow-up |
| Rauber(8) | UK adults | Quartiles of % total daily energy intake from UPF<br>24 hour recall measurement | Risk of 5% BMI increase<br>5 year median follow-up |
| Beslay(15) | French adults | Quartiles of % total daily grams intake from UPF<br>24 hour recall measurement | Risk of developing overweight or obesity<br>4.1 year median follow-up |
| Cordobra(16) | European adults | Quintiles of UPF grams per day<br>Combination of FFQ and 24 hour recall measurements | Risk of developing overweight or obesity<br>5 year follow-up |

Random effects meta-analysis using a Restricted Maximum Likelihood estimator were conducted using the ‘metafor’ package in R. Hazard Ratios were converted to Risk Ratios using the ‘toRR’ function from the ‘EValue’ package. We conducted separate meta-analyses to determine the effects of daily UPF intake on weight gain outcomes across different quartiles of UPF consumption (see table 1), with quartile 1 (lowest consumption of UPF) used as a comparator in each meta-analysis.

E values were calculated using the ‘evalues.RR’ function from the EValue package (see table 2). To convert Odds Ratios to Risk Ratios for the confounding effects we used the formula RR = OR / (1 – p0 + (p0 * OR)) where p0 is the baseline risk (6). To do this we used the ‘ORToRelRisk’ function from the ‘DescTools’ package (13). We used baseline risk estimates from relevant studies or conservative estimates if not available directly. Data and R code can be found here https://osf.io/z89pa/.

**Table 2.** Meta-analysis and corresponding E-values.

|  | Meta-analysis effect estimate (Risk Ratio) | E value for point estimate | E value for lower 95% confidence interval |
| --- | --- | --- | --- |
| Quartile 2 | 1.035 (1.006, 1.065) | 1.23 | 1.08 |
| Quartile 3 | 1.088 (1.041, 1.041) | 1.40 | 1.25 |
| Quartile 4 | 1.144 (1.100, 1.189) | 1.55 | 1.43 |

Lower and upper 95% confidence intervals in brackets. Reference category is quartile 1.

Table 2 presents the meta-analysis estimates and E values. Figure 2 illustrates the RR_UD_ and RR_EU_ values that equate to E values. Meta-analysis revealed statistically significant and small associations between UPF consumption and weight gain outcomes, based on effect size interpretation guidance (17). For the largest effect estimate (UPF quartile 4 vs. 1), E values for the point estimate and its lower confidence were 1.55 and 1.43, respectively, indicating that unmeasured confounders associated with both UPF consumption and weight gain to a similar degree (small to medium effect sizes) could nullify associations.

**Figure 2:**
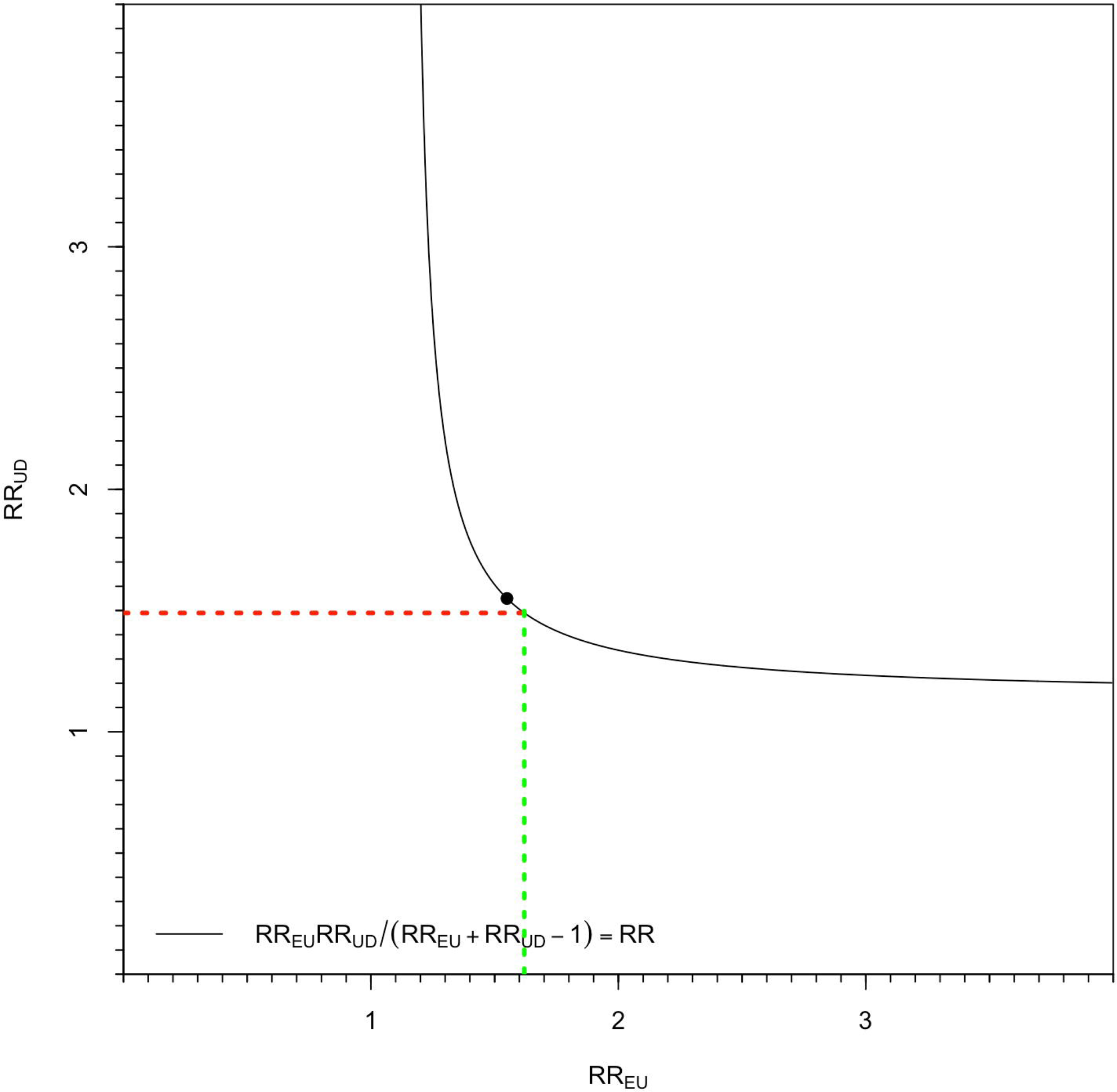
Plot of RR_UD_ and RR_EU_ and the E-Value for highest amount of UPF consumption association with weight gain. Red line is RR for overeating and weight gain, and green line represents the RR between overeating and UPF consumption needed to fully explain the association between UPF consumption and weight gain.

### Potential unmeasured confounding

No studies controlled for mental health or depression symptoms as potential confounders. Meta-analysis indicates that higher depression symptoms are associated with UPF consumption(18) (OR = 1.44 ∼ RR = 1.39) and predict development of obesity(19) (OR = 1.58 ∼ RR = 1.48). The joint bounding factor (*RR_UD_ * RR_EU_ /* (*RR_UD_+RR_EU_−1)) =* 1.10, suggesting that depressive symptoms could account for 71% (effect estimate) or as much as 100% (lower confidence interval of effect estimate) of the meta-analysed association between the highest vs. lowest UPF consumption groups and weight gain.

No studies controlled for eating traits, such as overeating. Tendency to overeat has a genetic basis and is typically characterised as either general disinhibited overeating or emotional-based overeating is associated with risk of higher BMI across multiple meta-analyses and effect sizes are medium in size (20, 21). For instance, the pooled association between disinhibited overeating and BMI is r = .28 (∼ OR = 2.88 [95% CI: 2.02 to 4.44] ∼ RR = 1.49 [95% CI: 1.34 to 1.63). Prospective studies of the association between tendency to overeat and weight gain produce similar estimates (22, 23). A positive relationship between tendency to overeat and higher UPF consumption would seem plausible, but there is a lack of data to confidently estimate effect size with precision and therefore concluded E-value calculation was not feasible. However, as denoted in Figure 2, an RR=1.62 (small to medium in size) would be needed to fully explain the meta-analysed association between the highest vs. lowest UPF consumption groups and weight gain.

### Potential residual confounding

All studies controlled for SES indicators, but not access to healthy nutritious food or food insecurity specifically. Food insecurity is associated with higher UPF consumption (24) and a recent epidemiological survey study estimates participants with the highest UPF consumption have a 60% higher prevalence of food insecurity (RR = 1.60)(25). Food insecurity is associated with elevated obesity risk in meta-analysis (OR = 1.53 ∼ RR 1.42) (26) and effects appear similar when examined prospectively (27). Taken together, it suggests that residual confounding of this nature could account for ∼86 % of the effect estimate, or 100 % based on the lower bound confidence interval.

## Discussion

We provide an illustration of how the E value can be used to examine the plausibility of confounding in obesity research. Using this approach we show that confounding variables with small-to-moderate sized associations with UPF consumption and weight gain could theoretically attenuate or completely nullify associations, and we identify a number of examples of potential confounding variables that could meet these conditions.

The present work also highlights some of the limitations of the E value. Accurate calculation is based on having sufficient statistical information about the potential confounders’ association with both outcome (RR_UD_) and exposure (RR_EU_) variables of interest. Overeating tendencies were identified as partially meeting the conditions for total confounding (RR_UD_ > lower bound E value confidence interval), but due to a lack of data to calculate a robust estimate RR_EU_, we were unable to formally apply the E value approach with confidence, though we were able to estimate associations between UPF consumption and overeating tendencies which would create statistical conditions that could nullify UPF-weight gain associations. This observation underscores that appropriate use of the E value is contextually specific and reliant on various effect size estimations, which may not always be available.

As the most extreme E values we identified were relatively small in size and a number of plausible confounders were identified that we reason could collectively attenuate the meta-analysed UPF consumption – weight gain association observed to non-significance, we propose that unmeasured confounding is of significant concern. Yet, it is important to note that the E value approach provides evidence on whether unaccounted confounding factors could *hypothetically* explain away observed associations. It is also plausible that more complete measurement of potential confounding variables could increase size of the UPF and weight gain association. This highlights the need for further confirmatory high quality observational research that is better able to control for potential confounders of concern and evidence from study types devoid of confounding (e.g., randomized controlled trials).

There is debate about how UPFs could causally contribute to weight gain. One proposed explanation is that the unfavourable macronutrient profile of UPFs could promote weight gain. A different, but not mutually exclusive explanation is that UPF consumption may also harm health independent to macronutrient profile and this is supported by some observational studies finding an association between UPFs and weight gain remains when macronutrient factors are controlled for(15). Meta-analysed studies did not consistently control for macronutrient profile. Therefore, from the present analyses it is unclear the extent to which the macronutrient profile of diets higher in UPFs could in part explain the meta-analysed associations observed and/or in combination with confounding variables, fully explain association between higher UPF consumption and weight gain.

There are limitations to the present research and the E value approach. We examined a select number of example potential confounders for illustrative purposes and other potential confounders may warrant investigation (e.g. shared genetic risk for weight gain and unhealthy diet). Food insecurity was examined as a quantifiable hypothetical example of residual confounding from measurement of SES. Most residual confounding by its nature is due to measurement imprecision and therefore unquantifiable. Food insecurity (based on prevalence) is a relatively rare event (dependent on country) and therefore itself may be unlikely to fully explain UPF and weight gain associations, but less extreme limited access to healthy nutrition will be more common and therefore a more likely potential confounder.

We based meta-analysis study inclusion on recent systematic reviews and not a formal search procedure, as this was beyond the scope of this technical report. A small number of studies were suitable for meta-analysis and they may be prone to publication bias. If so, the size of association between UPF consumption and weight gain and E-values may be overestimated.

The E value approach provides information on hypothetical confounding and the accuracy of estimates are dependent on the underlying statistical assumptions, as well as assumptions made when converting effect sizes. Critiques of the statistical application of the E value (6, 28) underscore that it can be at best considered as an analysis tool to inform thinking about potential confounding and not a blunt instrument to draw definitive conclusions from.

Unaccounted for confounding could plausibly explain the prospective association between UPF consumption and weight gain in adults. High quality observational research controlling for potential confounders and evidence from study types devoid of confounding are now needed.

## Data Availability

Data and R code can be found here https://osf.io/z89pa/.

## Contributions

Both authors conceived and designed the study. ER and AJ were responsible for data collection. AJ analysed the data. ER drafted the manuscript. Both authors revised the manuscript, contributed to the intellectual content and approved the final version.

